# High-dose high-intensity arm neurorehabilitation in chronic stroke improves general motor control

**DOI:** 10.1101/2025.11.18.25340491

**Authors:** Angelo C. Dawson, Joseph M. Galea, Sebastian Sporn, John W. Krakauer, Sven Bestmann, Nick S. Ward

## Abstract

**Background:** After stroke, patients can experience a rapid and generalised improvement in the control of movements, including movements that have not been trained (spontaneous recovery). This generalised improvement in motor control (or ‘behavioural repair’) is more effective in supporting functional recovery than task-specific training or behavioural compensation but has to date only been observed in the first few months after stroke.

**Methods:** We studied 81 chronic stroke patients at two time points, 3-weeks apart. 52 patients underwent a 3-week high-dose high-intensity upper limb neurorehabilitation programme (QSUL), and 29 chronic stroke patients receiving no treatment acted as control subjects (SC). At each time point, we assessed arm motor control kinematically using a 2D-planar reaching, a task which cannot be improved through compensation nor functional task training in 3D during neurorehabilitation. In addition, we measured strength, active range of joint motion and the upper extremity Fugl-Meyer score (FMA-UL).

**Results:** The FMA-UL increased by 6 points (IQR 3-8) in the QSUL-group and 0 points (IQR −1-1) in the SC-group. There were significant improvements in smoothness, movement time and accuracy of 2D-planar reaching in the QSUL-group compared to SC-group (all group x timepoint interactions P<0.03), which were independent of changes in strength or active range of joint motion.

**Conclusions:** Chronic stroke patients retain the capacity for generalised improvement in motor control in response to high-dose high-intensity neurorehabilitation. Normal or closer to normal motor control should remain a therapeutic target for improving arm paresis at all stages after stroke.

## Introduction

Stroke is the leading cause of long-term neurological disability worldwide.^1,2^ One in four adults will experience a stroke and two-thirds of these will be under the age of 70.^3^ Upper limb impairment is a major contributor to disability and occurs in approximately 75% of stroke patients.^4–6^ Improvement in upper limb motor function can come about in several ways. Firstly, because the upper limb has multiple degrees of freedom, stroke patients may simply achieve a task in a different way using alternative muscle groups. These compensatory strategies might lead to problematic secondary biomechanical changes in the limb. Furthermore, in primate models of stroke, behavioural compensation has a detrimental effect on the ability to achieve more normal patterns of upper limb movement through improved motor control.^7,8^ Secondly, improved performance on a motor task can be achieved through repetitive task-specific motor skill training (given the limits of individual impairments^9^). However, the gains in motor skill from practicing one task are not easily transferred or generalized to a different but related task.^10^ Thirdly, we can also observe a generalised, sometimes spontaneous, improvement in motor control often considered to represent true recovery or behavioural repair/restitution.^11,12^ Importantly, a generalised improvement in motor control can only be detected though an improvement in movement kinematics (not compensation) on a task that has not been trained (not task-specific training). Whilst compensation and task-specific motor improvements are available as strategies for functional improvement at any time after stroke, behavioural repair is thought to be dependent on specific molecular and cellular changes in the post-stroke brain that result from the expression of a ‘recovery transcriptome’^13^ only during the first few weeks or months after injury, and is often referred to as spontaneous biological recovery.^14^

Consequently, upper limb treatment in the chronic phase tends to focus on promoting behavioural compensation to improve an individual’s ability to participate in activities of daily living.^15^ Behavioural repair is not considered possible beyond the subacute post-stroke period. Recent work has investigated the effect of high-dose high-intensity upper limb neurorehabilitation that targets improving movement quality and control as well as achieving functional movement goals in chronic stroke patients.^16–19^ Large clinical improvements were seen in a range of standard outcome measures that assess both motor impairment and limitations of upper limb activity. These results have raised the possibility that a generalised improvement in motor control might after all be possible in chronic stroke patients in response to a high enough dose and intensity of neurorehabilitation. However, standard outcome measures used do not allow disambiguation of behavioural repair from compensation, task specific training, and improvements in strength.^20^ Demonstrating a generalised improvement in motor control requires (i) a kinematic assay of improved motor control (to exclude the possibility of compensation), and (ii) should be seen in motor tasks that have not been specifically trained (to exclude the effects of task-specific motor skill acquisition). We therefore investigated whether high-dose high-intensity upper limb neurorehabilitation could lead to generalised improvements in motor control in chronic stroke patients. If so, then behavioural repair should remain a therapeutic target well beyond the sub-acute stroke period.

## Methods

### Patient recruitment

We recruited two separate non-randomised groups of chronic (≥ 6 months from stroke onset) stroke patients with persistent upper limb motor impairment: (i) a cohort undergoing routine clinical treatment on the Queen Square Upper Limb neurorehabilitation programme (QSUL) between 1^st^ February 2017 and 1^st^ August 2019 were recruited (QSUL-group); (ii) a control group on the waiting list for QSUL who were not undergoing any treatment (SC-group) (Supplementary Figure S1). The control group was included to determine whether repeated testing on the kinematic assessment device itself could lead to changes in kinematic measures. Study inclusion criteria were age over 18-years; first ever stroke; no other brain injury, neurological condition or major psychiatric illness; no significant visual or hearing impairment, hemi-spatial neglect, severe dysphasia or musculoskeletal problem or pain limiting ability to participate in tasks or follow the study protocol.

### Treatment programme

QSUL is a single centre clinical service which provides 90 hours of timetabled treatment. A full description of the programme has been published using the TIDieR checklist in relation to a separate randomised control trial.^21^ QSUL aims at reducing impairment and promoting re-education of motor control within functional tasks. Individualised meaningful tasks were practiced repeatedly, to facilitate task mastery with a focus on quality of movement and minimisation of compensatory movements. Coaching was considered a key component of the programme, used throughout to embed new skills and knowledge into individual daily routines. Consequently, individuals increase participation and confidence in their desired goals, enhancing self-efficacy and motivation to sustain behavioural change beyond the end of the active treatment period.^22^

This overall approach was achieved through two daily sessions each of physiotherapy and occupational therapy, supplemented with tailored, individualised interventions, including repetitive practice with a rehabilitation assistant, sensory retraining, use of dynamic and functional orthoses, neuromuscular electrical stimulation, and group work. A 6-hour timetable was implemented 5 days a week for 3 weeks.

Patients in the QSUL-group were assessed at baseline (T1, day 1 of treatment) and 3 weeks later (T2, last day of treatment). Patients in the SC-group were assessed at baseline (T1) and again 3-weeks later (T2). At each timepoint, the Fugl-Meyer Assessment of the Upper Limb (FMA-UL),^23–25^ active elbow joint ROM, biceps/triceps strength and a range of kinematic measures were recorded. To test whether changes in kinematics are maintained once intensive upper limb treatment has ended, we followed up as many QSUL-group patients as possible at 6 weeks (T3) and 6 months (T4) post-discharge.

### Clinical assessment

The FMA-UL is a stroke-specific, performance-based index of motor impairment with good validity and intra- and inter-rater reliability. It tests whether patients can move the impaired upper limb without the intrusion of flexor synergistic patterns (a component of hemiparesis). The minimum clinically important difference (MCID) for FMA-UL is 5.25 points.^26^

Active ROM at the elbow was measured in line with standard methods using a double-armed protractor goniometer.^27^ Patients sat on a straight-backed chair with their torso supported against the back rest but no restriction to arm movement and the arm to be tested was held parallel to the midline of the body in the anatomical position. The number of degrees of movement from the starting position of the impaired and less impaired forearm to its position at the end of its full active ROM were recorded during elbow flexion and extension. If increased tone in the impaired arm limited active ROM, the examiner waited 2 minutes before measuring active ROM to allow the arm to relax as much as possible.

Muscle strength (maximum force of voluntary contraction, *kg*) in the arm during voluntary elbow flexion and extension was measured using a hand-held digital muscle dynamometer (microFET2™ System Hoggan Scientific, LLC. Capacity 300 lbs, calibrated to +/- 1% or 0.01 Kgf, Pro Med Products, Atlanta, GA) capable of measuring force in multiple planes. A standardised isometric muscle action procedure was used to test each movement,^28^ in line with methods used in similar recent work.^11^ Participants sat on a straight-backed chair with their torso supported against the back rest but no restriction to arm movement. Each strength trial lasted 7 seconds, comprising 2 seconds of building up to maximal strength and then 5 seconds sustaining maximal strength against the muscle dynamometer, followed by a 60 second rest period. Three strength trials were performed for each movement tested and the average peak force value was calculated. Strength trials and rest periods were timed by the examiner using a pre-programmed digital timer on an iPhone App (“Seconds” Version 3.18 (330) made by Apple® Runloop for IOS). This procedure was then repeated for the less impaired arm.

### Kinematic assessment

Arm motor control was assayed using a 2D-reaching task performed with a two-joint robotic manipulandum that removes the requirement for weight bearing by providing anti-gravity support (de-weighting) and allows near-frictionless movement in a horizontal plane via an air-sled system.^10,11,29^ This represents an assay for dexterous fractionation around the shoulder and elbow whilst adopting an unnatural posture with support against gravity as recommended by the International Stroke Recovery and Rehabilitation Alliance (ISRRA)^20^ for distinguishing between ‘true recovery’ and adoption of ‘compensatory’ movement. It is used because there is only one position in cartesian space per set of joint angles, so that compensation is not possible.

Patients sat in a high-backed chair in front of a workstation housing the manipulandum. A strap secured their trunk to the chair back and a headrest supported their forehead to minimise truncal compensatory movement and maximise comfort. Seat height was adjusted to bring the elbow and wrist into horizontal alignment with the handle of the manipulandum at mid-chest level. The forearm rested in a specially moulded cast secured with two straps, which fully supported the weight of the arm and splinted the wrist. The hand grasped the cylindrical handle of the manipulandum. A horizontal mirror suspended 15 cm above the position of the arm reflected a computer monitor mounted above it, on which the task was projected, and prevented direct vision of the working arm (Figure 1). The position of the cylindrical handle (corresponding to hand position) was represented by a cursor on the monitor.

**Figure 1.**
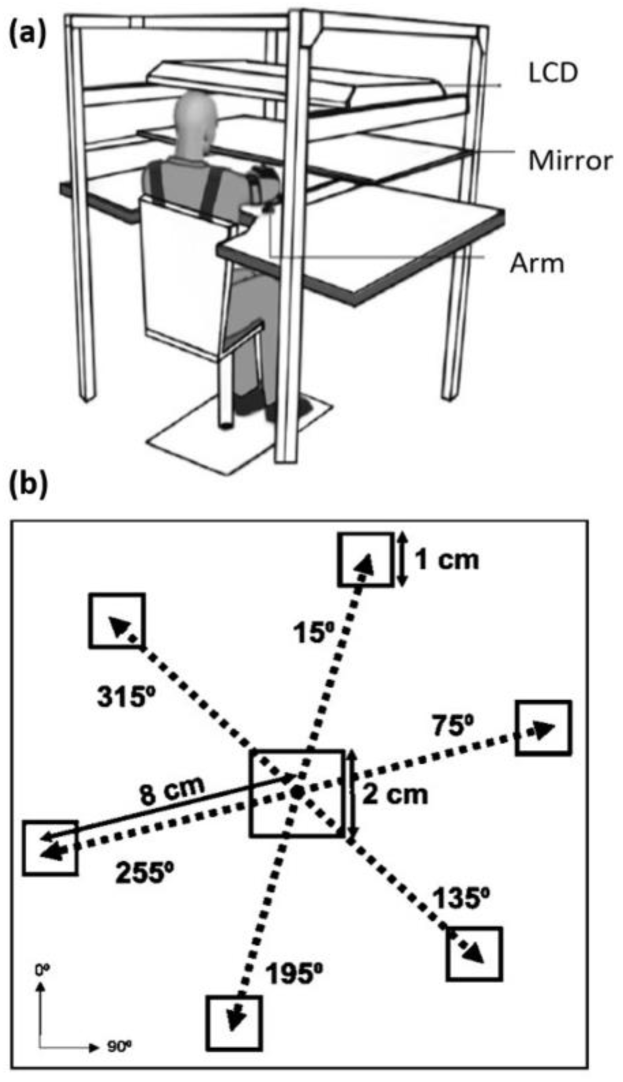
Experimental setup for measuring motor control. a) Schematic diagram of the experimental set-up. A horizontal mirror suspended 15 cm above the position of the arm reflected a computer monitor (LCD) mounted above it, on which the task was projected, and prevented direct vision of the working arm (Diagram adapted from Cortes et al., 2017).^11^ b) Position of target angles in relation to the patient when the right arm is performing the task. Targets at 135°, 195° and 255° require elbow flexion (movement within flexor synergy). Targets at 15°, 75° and 315° require elbow extension (movement against upper limb flexor synergy).

Patients made straight-line movements with the cursor from a central starting square (2 cm^2^) towards radially arranged target squares (1 cm^2^) that appeared in a pseudo-random order at 8 cm distance from the centre of the start square, equally spaced at 15°, 75°, 135°, 195°, 255° and 315° with reference to the participant’s sagittal midline (Figure 1b). Patients had 3 seconds in which to complete each trial and were instructed to move as quickly and accurately as possible. One target square appeared per trial with each target angle appearing 5 times during a block of 30 trials. If a movement successfully ended inside the target with a peak speed of 0.01 to 0.40 m/s, the target square turned green and “exploded” in a visually pleasing manner. If a movement did not successfully end inside the target within the 3 second timeframe or achieve the required peak speed, the target remained white. Regardless of the position of the cursor, the target square disappeared 3 seconds after its appearance and the robot returned the hand to the starting position, signalling the end of each trial.

Patients completed one block of 18 practice trials with their less-impaired arm followed by one block of 18 practice trials with their impaired arm to gain familiarity with the task. After a two-minute rest period, one block of 30 recorded trials was performed with the less-impaired arm followed by one block of 30 recorded trials with the impaired arm. Following a further two-minute rest period, the final block of 30 recorded trials was performed by the less-impaired arm followed by the impaired arm to minimise fatigue.

The 2D (x, y) position of the hand in the robotic manipulandum was collected through a custom C++ code at a pre-defined sampling rate of 200 Hz. Derivation of kinematic measures, data science and statistical analysis were subsequently performed ‘offline’ using Matlab (version R2017b, The MathWorks, Natick, MA, USA) and R Studio (R Core Team (2019) R: A language and environment for statistical computing. R Foundation for Statistical Computing, Vienna, Austria. URL https://www.R-project.org/).

To decrease contamination from target guesses, incomplete movements and unintentional movements, trials were excluded for the purposes of subsequent analysis when movement length ≤ 4cm; maximum speed < 0.06 m/s and direction at peak speed was ≥ 90⁰ away from the target angle, in line with previous work^11^. All other trials were included for subsequent analysis.

Movement-start was defined as the time before peak speed when speed > 0.02 m/s; movement-end was defined as time after peak speed when speed ≤ 0.02 m/s for > 0.10 sec. Movement time was defined as the time between movement onset and movement end (ms). Peak speed (also referred to as maximum speed per trial) was defined as the first zero-crossing of acceleration when speed > 0.01 m/s. Maximum velocity was defined as the maximum velocity (m/s) between movement start and movement end.

To measure movement accuracy, radial distance from target was calculated for successful trials, taking the radial (√ (x^2^ + y^2^)) distance to the centre of the target square when velocity was 0.01 m/s and movement had stopped for longer than 40 ms. Successful trials were defined as those where the end position was < 0.5cm from the target edge when the speed < 0.01 m/s for at least 40 ms, within 3.0 seconds.

To measure movement smoothness, we calculated the log dimensionless jerk from the logarithm naturalis of the sum of the squared acceleration multiplied by the trial duration to the power of three and divided by the squared peak velocity. Data were first low-passed filtered.

In addition, the primary outward submovement for each whole movement trajectory was defined as the initial movement segment up until the first minimum velocity (<0.005 m/s). The duration of the primary outward submovement as a proportion of the duration of the whole movement trajectory, primary outward submovement log dimensionless jerk (movement smoothness) and primary outward submovement radial distance from target (movement accuracy) were then calculated as described above.

### Analysis Plan

1. All single time point data were represented by medians and interquartile ranges (IQRs) for transparency. We undertook a planned comparison of T1 to T2 changes in kinematic and clinical measures for the more impaired (‘impaired’) arm between QSUL and SC-groups. Our primary measures of interest were movement kinematics (i.e. movement smoothness, movement accuracy, movement time). Our secondary measures were FMA-UL, active elbow ROM, and muscle strength. Differences in T1-T2 change scores between QSUL-group and SC-groups were compared using 2x2 mixed ANOVAs. Subsequent confirmatory post-hoc 2-sample 2-tailed t-tests comparing the mean T1-T2 changes for QSUL versus mean T1-T2 changes for SC-groups were conducted to reveal direction of change, adjusting P values for multiple comparisons using the Bonferroni method.
2. Additional control analyses were performed to determine if kinematic changes (i) reflected a change in speed-accuracy trade off; (ii) were independent of active elbow ROM and elbow strength; (iii) were due to improved movement execution or feedback control (online corrections); (iv) were simply due to online learning during kinematic analysis (Supplementary material).
3. To test whether repeated testing on the 2D-planar device did not itself lead to an improvement in kinematics through learning, we compared T1-T2 change scores between (i) the less impaired arm for the QSUL-group and the SC-group, and (ii) the more impaired and less impaired arm in the QSUL-group. (Supplementary material).
4. To test whether changes in kinematics are maintained after treatment, changes in FMA-UL, active elbow ROM and strength and kinematic measures were compared between T1-T2, T1-T3 and T1-T4 using one-way ANOVAs and subsequent multiple pair-wise comparisons using the Bonferroni method to adjust P values for multiple comparisons.

All statistical tests were performed in R Studio (version 3.6.1 (2019-07-05)).

### Ethical Approval

This study was reviewed and approved by the London Camden and Kings Cross Research Ethics Committee (REC reference 17/LO/1466) and conducted in accordance with the Declaration of Helsinki (2013).

## Results

### Study population

We recruited 81 chronic stroke patients (52 in QSUL-group; 29 in SC-group) who were assessed at two time points 3-weeks apart (T1 and T2). The clinical and demographic characteristics of each group were well-matched (Table 1) and although there was a significant difference in time since stroke onset (QSUL-group 24.8 (13.2) months; SC-group 40.6 (23.1) months, *P* = 0.74 x 10^-3^), both study groups met the definition of chronic stroke.^30^ We had no access to information on whether patients had ischaemic or haemorrhagic stroke, but in the chronic stage of stroke, this has no effect on subsequent response to motor rehabilitation beyond the clinical deficit. Patient recruitment and exclusions are shown in Figure S1.

**Table 1.** Baseline clinical and demographic characteristics of QSUL- and SC-groups at baseline (0 weeks)

| Characteristic | QSUL-group (n = 52) | SC-group (n = 29) |
| --- | --- | --- |
| Gender – male: female | 25:27 | 15:14 |
| Handedness (prior to stroke) – right: left | 47:5 | 25:4 |
| Affected upper limb – dominant: non-dominant | 25:27 | 13:16 |
| Age (years), mean (SD) | 52.8 (16.1) | 56.0 (11.0) |
| Time since stroke (months), mean (SD) | 24.8 (13.2) | 40.6 (23.1) |
| Fugl Meyer Assessment Upper Limb (FMA-UL) (total 54 points), mean (SD) | 24.3 (8.8) | 24.5 (11.6) |
| Modified Rankin Score (MRS), mean (SD) | 2.7 (0.6) | 2.7 (0.6) |

### Clinical scores

The median change in FMA-UL scores from T1 to T2 was 6 points (interquartile range (IQR) 3-8) in the QSUL-group and 0 points (IQR −1-1) in the SC-group (Figure 2). We observed a group x timepoint interaction for FMA-UL (*F*(1,73) = 2.43, *P* = 0.42 x 10^-11^) indicating greater increase in the QSUL-group compared to the SC-group (Table 2).

**Figure 2.**
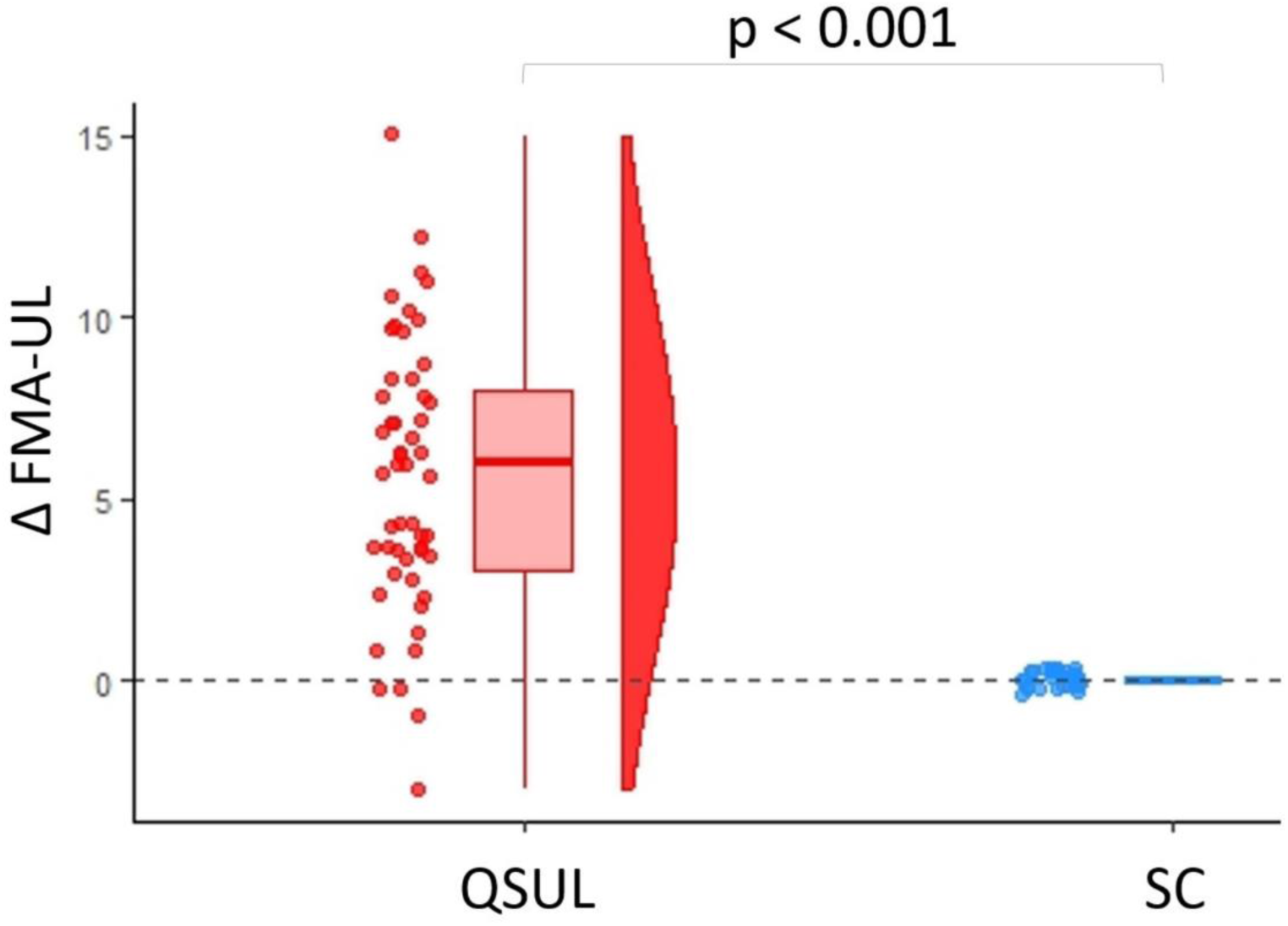
Clinical measures improved following upper limb treatment. Change (Δ) in impaired arm FMA-UL score in the QSUL-group (red) compared to the SC-group (blue). A more positive change score is better. Changes in FMA-UL between T1 and T2 in the QSUL-group and SC-group were compared using 2x2 mixed ANOVAs to test for significant group x timepoint interactions (Table 2) and subsequent post-hoc 2-sample t-tests to reveal direction of change (all P values < 0.05 after correction for multiple comparisons. using the Bonferroni method).

**Table 2.** Change (Δ) in measures between T1 and T2 for the impaired arm of the QSUL-group (n = 52) compared to the impaired arm of the SC-group (n = 29)

| Change ($\Delta$) T1-T2 | QSUL-group | SC-group | ANOVA<sup>a</sup> | |
| --- | --- | --- | --- | --- |
|  | Med (I/4-3/4) | Med (I/4-3/4) | F(1,73) | p |
| **Clinical Scores** |  |  |  |  |
| $\Delta$_FMA-UL (points) | 6.0 (3-8) | 0.0 (-1-0) | 2.43 | $0.42 \times 10^{-11}$ |
| **Clinical Measures** |  |  |  |  |
| $\Delta$_Biceps strength (kgf) | 0.8 (0.2-2.1) | 0.0 (-0.2-0.2) | 1.50 | $0.23 \times 10^{-4}$ |
| $\Delta$_Triceps strength (kgf) | 0.6 (0.2-2.4) | -0.2 (-0.4-0.2) | 1.60 | $0.11 \times 10^{-3}$ |
| $\Delta$_Elbow flexion (degrees) | 2.0 (0.0-5.0) | 0.0 (0.0-1.0) | 8.12 | 0.01 |
| $\Delta$_Elbow extension (degrees) | 0.0 (-9.6-0.0) | 0.0 (-2.0-0.0) | 4.50 | $0.13 \times 10^{-2}$ |
| **Kinematic measures** |  |  |  |  |
| $\Delta$_Movement smoothness (jerk) | -0.7 (-1.4-(-0.1)) | -0.2 (-0.5-0.0) | 2.10 | 0.02 |
| $\Delta$_Movement accuracy (cm) | -0.02 (-0.05-0.00) | 0.00 (-0.02-0.03) | 3.31 | 0.03 |
| $\Delta$_Movement time (ms) | -227 (-456-10) | -90 (-168-(-15)) | 2.73 | 0.01 |

<sup>a</sup>Changes in clinical scores, clinical measures and kinematic measures between T1 and T2 in the QSUL-group and SC-group were compared using 2x2 mixed ANOVAs to test for group x timepoint interactions Med: median value, I/4: 25<sup>th</sup> percentile, 3/4: 75<sup>th</sup> percentile.

The median change in active elbow extension ROM from T1 to T2 was 0.0 (IQR −9.25-0.0) degrees in the QSUL-group and 0.0 (IQR −2.0-0.0) degrees in the SC-group. There was a group x timepoint interaction *F*(1,73) = 4.5, *P* = 0.13 x 10^-2^ (Table 2). The median change in active elbow flexion ROM from T1 to T2 was 2.00 (IQR 0.00-5.00) degrees in the QSUL-group and 0.0 (IQR 0.0-1.0) degrees in the SC-group. There was a group x timepoint interaction *F*(1,73) = 8.12, *P* = 0.01 (Table 2).

The median change in biceps strength from T1 to T2 was 0.8 (IQR 0.2-2.1) kgf in the QSUL-group and 0.0 (IQR −0.2-0.2) kgf in the SC-group. There was a group x timepoint interaction *F*(1,73) = 1.5, *P* = 0.23 x 10^-4^ (Table 2). The median change in triceps strength from T1 to T2 was 0.6 (IQR 0.2-2.4) kgf in the QSUL-group and −2.0 (IQR −0.4-0.2) kgf in the SC-group. There was a group x timepoint interaction *F*(1,73) = 1.6, *P* = 0.11 x 10^-3^ (Table 2).

Post-hoc 2-sample t-tests confirmed significant improvements between T1 and T2 in the QSUL-group impaired arm compared to the SC-group impaired arm for FMA-UL, biceps and triceps strength, flexion and extension active ROM (all *P* < 0.05, corrected for multiple comparisons.

### Motor control (Kinematic measures)

The median change in movement smoothness (logdimjerk) from T1 to T2 was −0.68 (IQR - 1.36-(-0.11) (representing reduced jerk and therefore improvement in smoothness) in the QSUL-group and −0.2 (IQR −0.5-0.0) in the SC-group. There was a group x timepoint interaction (*F*(1,73) = 2.1, *P* = 0.02) (Table 2, Figure 3a).

**Figure 3.**
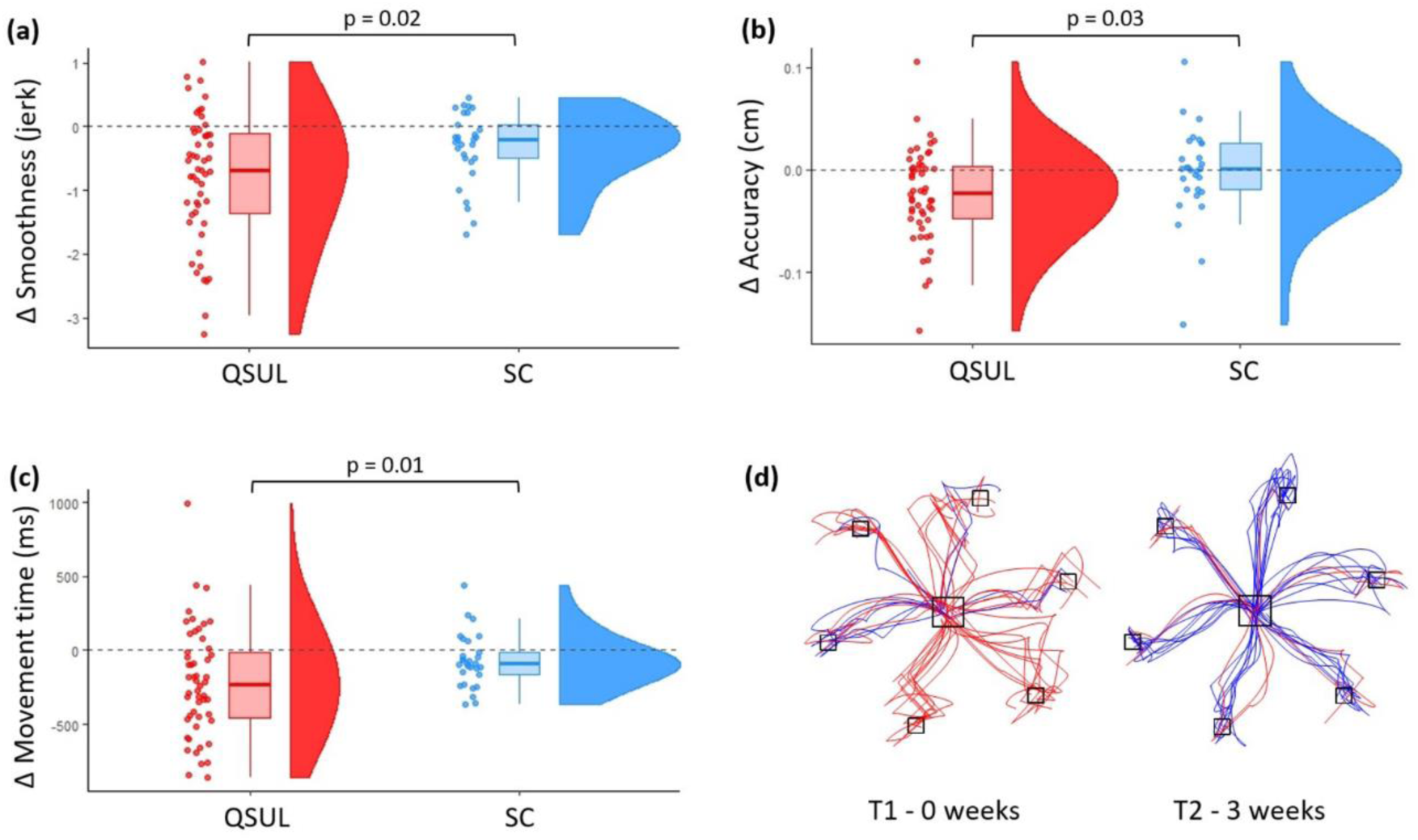
Kinematic measures improved following upper limb treatment. a-c) Change in key kinematic parameters (movement smoothness (jerk), movement accuracy (cm), movement time (ms)) of whole movement trajectories performed by the impaired arm in the QSUL-group compared to the impaired arm in the SC-group. The dashed grey line represents no change. The boxplots denote the median and interquartile range values. A more negative change score is better. Changes in kinematic measures between T1 and T2 in the QSUL-group and SC-group were compared using 2x2 mixed ANOVAs to test for significant group x timepoint interactions (Table 2) and subsequent post-hoc 2-sample t-tests to reveal direction of change (all P values < 0.05 after correction for multiple comparisons. using the Bonferroni method). d) Example of impaired arm reaching movements made by a QSUL-group patient at T1 and T2 over the 32-week study period. Unsuccessful trials in red, successful trials in blue.

The median change in movement accuracy (radial distance from target) from T1 to T2 was - 0.02 (IQR 0.05-0.0) cm (representing movements that were closer to the target) in the QSUL-group and 0.0 (IQR −0.02-0.03) cm in the SC-group. There was a group x timepoint interaction *F*(1,73) = 3.3, *P* = 0.03 (Table 2, Figure 3b). Note, movement accuracy analysis was only performed on successful trials, defined as those where the end position was < 0.5cm from the target edge when the speed < 0.01 m/s for at least 40 ms, within 3.0 seconds. For this reason, the absolute differences in movement accuracy appear small, but were nevertheless significantly improved in the QSUL group compared to SC-group.

The median change in movement time from T1 to T2 was −227 (IQR -455-10) ms in the QSUL-group and -90 (IQR −168-(-15)) ms in the SC-group. There was a group x timepoint interaction *F*(1,73) = 2.7, *P* = 0.01 (Table 2, Figure 3c).

Post-hoc 2-sample t-tests confirmed significant improvements between T1 and T2 in the QSUL-group impaired arm compared to the SC-group impaired arm for movement smoothness, movement accuracy and movement time (all *P* < 0.05, corrected for multiple comparisons).

Additional control analyses demonstrated that kinematic changes (i) reflected a change in speed-accuracy trade off (Figure S2, Figure S3); (ii) were independent of active elbow ROM and elbow strength (Table S1); (iii) were due to improved movement execution rather than feedback control (online corrections) (Figure S4); (iv) were not due to online learning during kinematic analysis (Figure S5).

Additional control analyses involving the less impaired arm of each group found that (i) in the QSUL-group, changes in impaired arm were greater than those for less impaired arm (Table S2, Figure S6), and (ii) there was no significant change in the less impaired arm in either QSUL-group or SC-group, apart from small relative improvement in biceps and triceps strength of QSUL less impaired arm compared to SC-less impaired arm (Table S3, Figure S6). Taken together, these results suggest that repeated testing on the 2D-planar device did not itself appear to lead to an improvement in kinematics through learning, and so the changes we report for the impaired arm were unlikely to be driven by the repeated testing itself.

Data were also collected at 6 weeks (T3) and 6 months (T4) after treatment was finished in 28 participants from the QSUL-group only. A significant effect across timepoints (T1, T2, T3 and T4) was seen for all clinical and kinematic measures (Table 3, Figure 4). The pairwise comparison for T1 vs T2, T1 vs T3, and T1 vs T4 were significant for all clinical and kinematic measures (Table 3). Overall, the significant gains in clinical and kinematic measures seen at the end of the neurorehabilitation intervention (T2) were maintained at T3 and T4.

**Figure 4.**
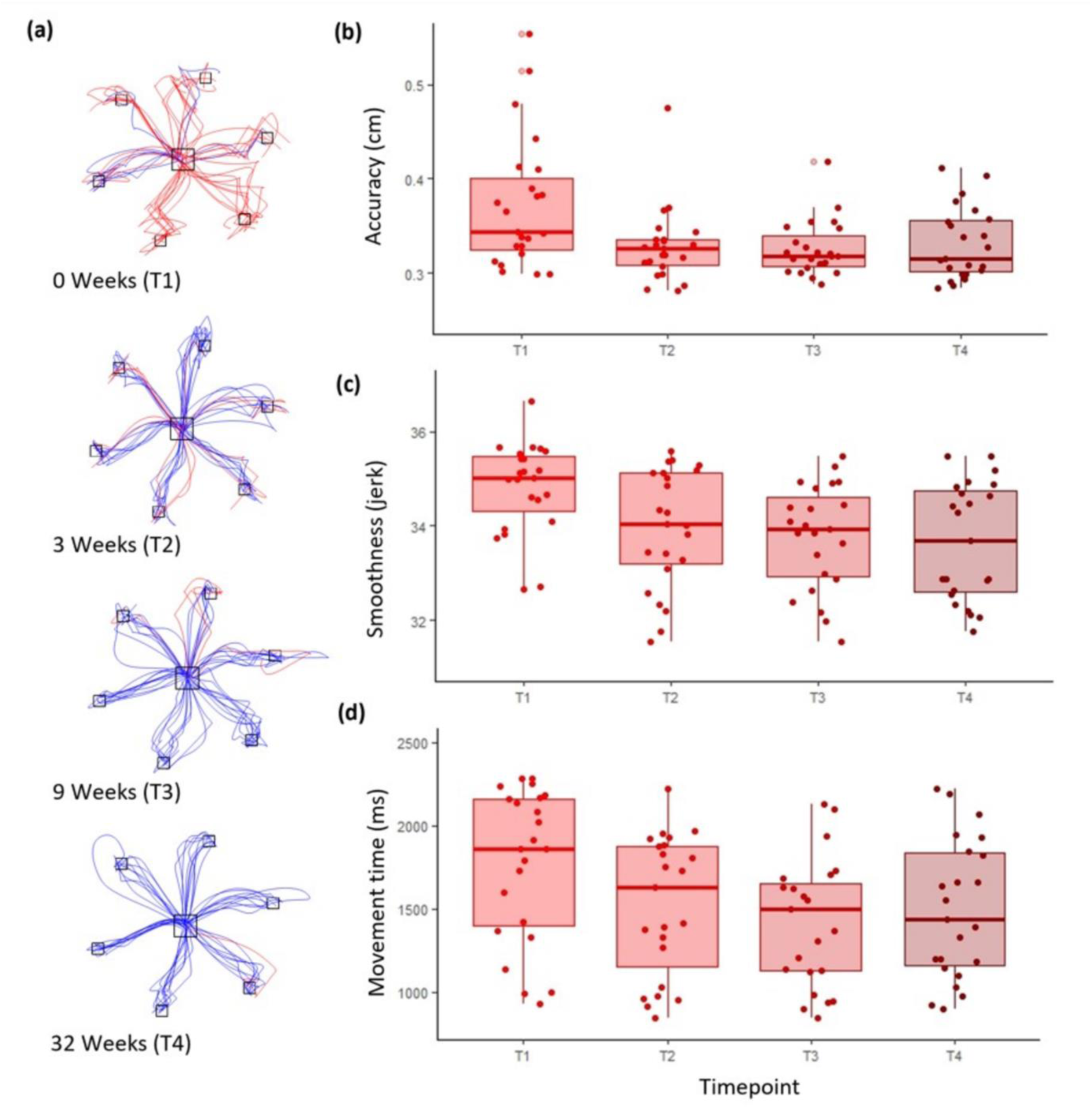
Improvements in motor control in the QSUL-group were retained beyond the end of treatment. **a)** Example of impaired arm reaching movements made by a QSUL-group patient over the 32-week study period. Unsuccessful trials in red, successful trials in blue. **b-d)** Improvement in key kinematic measures for whole movement trajectories performed by the impaired arm in the QSUL-group over the 32-week study period (n = 28). The boxplots denote the median and interquartile range values. Results of one-way repeated measures ANOVA tests to demonstrate an effect of time and pair wise comparisons for T1 vs T2, T1 vs T3 and T1 vs T4 are shown in Table 3.

**Table 3.**
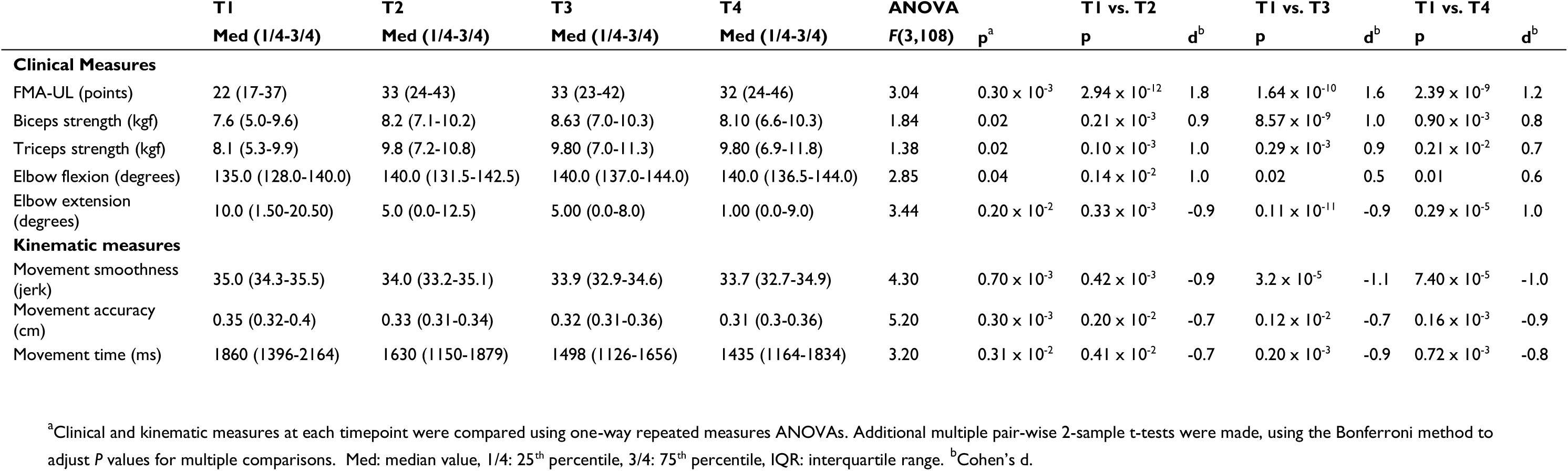
Clinical and kinematic measures at each timepoint (T1-T4) for the impaired arm of the QSUL-group (n=28)

|  | **T1** | **T2** | **T3** | **T4** | **ANOVA** |  | **T1 vs. T2** |  | **T1 vs. T3** |  | **T1 vs. T4** |  |
| --- | --- | --- | --- | --- | --- | --- | --- | --- | --- | --- | --- | --- |
|  | **Med (I/4-3/4)** | **Med (I/4-3/4)** | **Med (I/4-3/4)** | **Med (I/4-3/4)** | **F(3,108)** | **p<sup>a</sup>** | **p** | **d<sup>b</sup>** | **p** | **d<sup>b</sup>** | **p** | **d<sup>b</sup>** |
| **Clinical Measures** |  |  |  |  |  |  |  |  |  |  |  |  |
| FMA-UL (points) | 22 (17-37) | 33 (24-43) | 33 (23-42) | 32 (24-46) | 3.04 | 0.30 × 10<sup>-3</sup> | 2.94 × 10<sup>-12</sup> | 1.8 | 1.64 × 10<sup>-10</sup> | 1.6 | 2.39 × 10<sup>-9</sup> | 1.2 |
| Biceps strength (kgf) | 7.6 (5.0-9.6) | 8.2 (7.1-10.2) | 8.63 (7.0-10.3) | 8.10 (6.6-10.3) | 1.84 | 0.02 | 0.21 × 10<sup>-3</sup> | 0.9 | 8.57 × 10<sup>-9</sup> | 1.0 | 0.90 × 10<sup>-3</sup> | 0.8 |
| Triceps strength (kgf) | 8.1 (5.3-9.9) | 9.8 (7.2-10.8) | 9.80 (7.0-11.3) | 9.80 (6.9-11.8) | 1.38 | 0.02 | 0.10 × 10<sup>-3</sup> | 1.0 | 0.29 × 10<sup>-3</sup> | 0.9 | 0.21 × 10<sup>-2</sup> | 0.7 |
| Elbow flexion (degrees) | 135.0 (128.0-140.0) | 140.0 (131.5-142.5) | 140.0 (137.0-144.0) | 140.0 (136.5-144.0) | 2.85 | 0.04 | 0.14 × 10<sup>-2</sup> | 1.0 | 0.02 | 0.5 | 0.01 | 0.6 |
| Elbow extension (degrees) | 10.0 (1.50-20.50) | 5.0 (0.0-12.5) | 5.00 (0.0-8.0) | 1.00 (0.0-9.0) | 3.44 | 0.20 × 10<sup>-2</sup> | 0.33 × 10<sup>-3</sup> | -0.9 | 0.11 × 10<sup>-11</sup> | -0.9 | 0.29 × 10<sup>-5</sup> | 1.0 |
| **Kinematic measures** |  |  |  |  |  |  |  |  |  |  |  |  |
| Movement smoothness (jerk) | 35.0 (34.3-35.5) | 34.0 (33.2-35.1) | 33.9 (32.9-34.6) | 33.7 (32.7-34.9) | 4.30 | 0.70 × 10<sup>-3</sup> | 0.42 × 10<sup>-3</sup> | -0.9 | 3.2 × 10<sup>-5</sup> | -1.1 | 7.40 × 10<sup>-5</sup> | -1.0 |
| Movement accuracy (cm) | 0.35 (0.32-0.4) | 0.33 (0.31-0.34) | 0.32 (0.31-0.36) | 0.31 (0.3-0.36) | 5.20 | 0.30 × 10<sup>-3</sup> | 0.20 × 10<sup>-2</sup> | -0.7 | 0.12 × 10<sup>-2</sup> | -0.7 | 0.16 × 10<sup>-3</sup> | -0.9 |
| Movement time (ms) | 1860 (1396-2164) | 1630 (1150-1879) | 1498 (1126-1656) | 1435 (1164-1834) | 3.20 | 0.31 × 10<sup>-2</sup> | 0.41 × 10<sup>-2</sup> | -0.7 | 0.20 × 10<sup>-3</sup> | -0.9 | 0.72 × 10<sup>-3</sup> | -0.8 |

<sup>a</sup>Clinical and kinematic measures at each timepoint were compared using one-way repeated measures ANOVAs. Additional multiple pair-wise 2-sample t-tests were made, using the Bonferroni method to adjust *P* values for multiple comparisons. Med: median value, I/4: 25<sup>th</sup> percentile, 3/4: 75<sup>th</sup> percentile, IQR: interquartile range. <sup>b</sup>Cohen's *d*.

## Discussion

We have shown that high-dose high-intensity upper limb neurorehabilitation in the chronic phase of stroke can result in improvements of untrained 2D-planar arm movement trajectories. Crucially, these improvements did not result from behavioural compensation or from task-specific training effects (in 2D-planar movements) and were independent of either changes in active elbow range of movement or arm strength. Our results demonstrate that given the right conditions, a general improvement in arm motor control (i.e. behavioural repair) is possible in chronic stroke patients.

There is increasing acceptance that with a high enough dose of rehabilitation, motor performance can improve in the chronic phase of stroke,^19,31^ but the mechanisms of improvement are unclear. There is currently no evidence that neurorehabilitation can result in behavioural repair in chronic stroke, possibly because standard outcome measures have not allowed disambiguation of behavioural repair from either compensation or task specific training effects.^20^ A generalised improvement in motor control can only be detected where compensation and task-specific training effects can be ruled out.^11,12^ In our experimental motor control assay, compensation was not possible because there is only one position in cartesian space per set of joint angles. Furthermore, to avoid interpretation of our results as simple task-specific training effects, our motor control assay is deliberately far removed from any trained movements on the QSUL programme. During kinematic assessment, movement is made with the shoulder abducted (to approx. 80 degrees – not something trained for achieving function), elbow flexed, palm down, with no focus on wrist/fingers, and movement in 6 directions (Figure 1). Training of reaching during neurorehabilitation on the other hand is in naturalistic 3D, starting with the shoulder adducted, elbow flexed, forearm in mid-pronation with focus on hand shaping in relation to the object, especially wrist and finger extension and thumb abduction. Movement is always in a forward arc in relation to the body. We have taken advantage of the ’curse of task specificity’ in motor skill acquisition and carefully selected a motor control assay which does not overlap with training tasks to ensure that changes in kinematics were highly unlikely to be due to compensation or task-specific training effects.

Disambiguation of motor behavioural repair on the one hand, and compensation and task-specific training effects on the other, can be difficult.^20^ In one study, constraint-induced movement therapy led to improvements in the Action Research Arm Test but not arm kinematics suggesting any therapeutic effect was largely due to improved compensatory movement strategies.^29^ Conversely, robotic training of the impaired arm in chronic stroke patients led to improvements in arm kinematics but not clinical scores.^10^ In this case, a generalised improvement in motor control could not be inferred because patients were trained and tested on the same robotic device, and so arm kinematics likely improved through task-specific skill acquisition albeit without clinical improvement. One study did demonstrate that training in a naturalistic motor task (spooning beans from one cup to another) led to improvements in two untrained test tasks;^32^ (i) sorting wooden blocks from one box to another, and (ii) fastening buttons. It is important to note that both test tasks were assessed by measuring task outcome (time to completion), rather than quality of movement (kinematics). Naturalistic actions require many degrees of freedom and so improvement in task outcome may have been achieved through compensatory movements. In addition, similarities between training and one of the test tasks could result in task-specific training effects. Once again, despite the importance of this result for motor rehabilitation, one cannot infer a generalised improvement in motor control without assessing movement quality (kinematics).^20^

In our study, patients underwent what is considered from a qualitative perspective to be standard physical and occupational therapy, albeit at high-dose and intensity. Any robotic devices used for treatment operated in 3D-space and the time on these tasks was well below 5% of the therapy time.^16^ As discussed, we selected our 2D-planar reaching kinematic assessment task to be as independent from the training received as possible. Participants did perform a number of trials using the 2D-planar reaching device, which could have constituted training, but the less impaired arm of our patients and the impaired arm of control patients performed the same number of trials without resultant changes in kinematics. There is no suggestion that the kinematic improvements were due to making slower movements. In fact, our kinematic results suggest a large improvement in movement time in the QSUL group together with a small (but statistically significant) improvement in accuracy. This is reflected in an improved speed-accuracy trade off function in the QSUL group (Supplementary Figure S2, S3). In addition, further analysis did not show any online learning effects in either kinematic testing session (Supplementary Figure S5). It is also notable that changes in clinical scores, active elbow ROM and arm strength, were all independent of changes in arm kinematics, highlighting the point that the phenotype of hemiparesis is made up of several separate components that respond independently to different aspects of neurorehabilitation. Together, our results provide the most compelling evidence to date that a general improvement in motor control, or behavioural repair is possible in the chronic stage of stroke.

One possible criticism is that our kinematic changes may not be considered clinically meaningful. However, the key point here is that our results are an important proof of principle. We have identified a credible biological signal for true improvement in motor control, previously thought not to exist. This signal can now become a target for larger effect sizes through new forms of behavioural intervention and their augmentation by physiological stimulation. We must also grasp a point that the field of neurorehabilitation research has consistently failed to understand; that a seemingly small change in one measure may create the optimal conditions for other treatments to work. As an example, although re-establishing 10 degrees of voluntary finger extension may not by itself have an impact on daily function, it opens the door for other treatments (e.g. constraint induced movement therapy) to achieve clinically meaningful change.

How might our treatment programme accomplish restoration of motor control? Although our data are the first to plausibly demonstrate behavioural repair through neurorehabilitation in chronic stroke patients, other recent work has shown a generalised improvement in motor control in chronic stroke patients in response, not to training, but to epidural cervical spinal cord stimulation.^33^ Turning on the epidural stimulator was thought to increase excitability of the spinal cord motoneurons and led to an instantaneous improvement in both strength and untrained motor control (assessed as in our study with 2D-planar reaching kinematics). The speed with which behavioural improvements were gained and then lost when the device was turned on then off suggests that there was a residual latent motor system capacity that could rapidly be brought online to support behavioural repair at the physiological level. The latent motor system is most likely to be the corticospinal rather than reticulospinal system, since the latter is unlikely to support improvement of out-of-flexor-synergy motor control.^34^ In other words, shifting of the excitatory threshold at the spinal cord motoneuron might allow a latent but functioning corticospinal system to exert immediate control over strength and motor control. Importantly, at least in the case of epidural stimulation for gait recovery after spinal cord injury, the effect of epidural stimulation is greatly amplified by training.^35^ In this scenario, a generalised improvement in motor control might be the consequence of directed motor practice layered on top of this unmasked corticospinal system capacity.

One explanation for our results then, is that in a subset of patients, very high-dose high-intensity neurorehabilitation can unmask latent corticospinal system capacity and achieve a generalised improvement in motor control through training, something that currently recommended pragmatic task-based upper limb compensation does not achieve. Training itself may not achieve behavioural repair^9^ unless the right neural conditions are present. Here the ‘right neural conditions’, based on work in animal models of stroke, are those expressed during spontaneous biological recovery.^36–38^ There are important parallels in pre-clinical models of stroke, where the combination of motor training and environmental enrichment, influence many of the molecular changes associated with spontaneous biological recovery. Animals undergoing enriched rehabilitation exhibited recovery of reaching behaviour through improved motor control and a reduction in abnormal motor synergies.^39–41^ One possibility then is that our patients have been exposed to conditions which overlap mechanistically with spontaneous biological recovery, thereby increasing the possibility of a generalised improvement in motor control,^42,43^ but we have no way of confirming this in our study. Future work will need to both identify those patients with latent recovery capacity and optimize intervention approaches, whether enriched behavioural training, physiological or pharmacological, that bring this capacity online.

We present new evidence that a general improvement in motor control is achievable in the chronic phase of stroke with high-dose high-intensity upper limb training. Improving general motor control should therefore remain a therapeutic target in the chronic stage of hemiparetic stroke because this will support improved recovery and better quality of life. We must now explore how to maximise improvements in chronic stage general motor control by identifying new forms of motor training that optimize for limb control that go beyond simply increasing the dose of regular upper limb neurorehabilitation (recommended to be repetitive task training in most countries). Our results also re-emphasise that the phenotype of hemiparetic impairment is made up of several distinct and independent impairments.

Ultimately, optimising stroke motor recovery is likely to require a pluralistic training approach that differentially targets the components of hemiparetic impairment, as well as the different levels of the International Classification of Functioning, Disability and Health (ICF).

### Data availability

The data will be shared upon reasonable request to the corresponding author. This study did not generate any large datasets.

## Acknowledgements

We would like to acknowledge all the staff and participants from the Queen Square Upper Limb Neurorehabilitation programme; Mr Paul Hammond for expert technical support in setting up and maintaining the kinematics lab.

## Funding

SS is funded by the Jon Moulton Charity Trust.

### Supplementary material

Supplementary material is available online.

