## Supplemental material for "High-dose high-intensity arm neurorehabilitation in chronic stroke improves general motor control"

### Supplementary material: Consort diagram

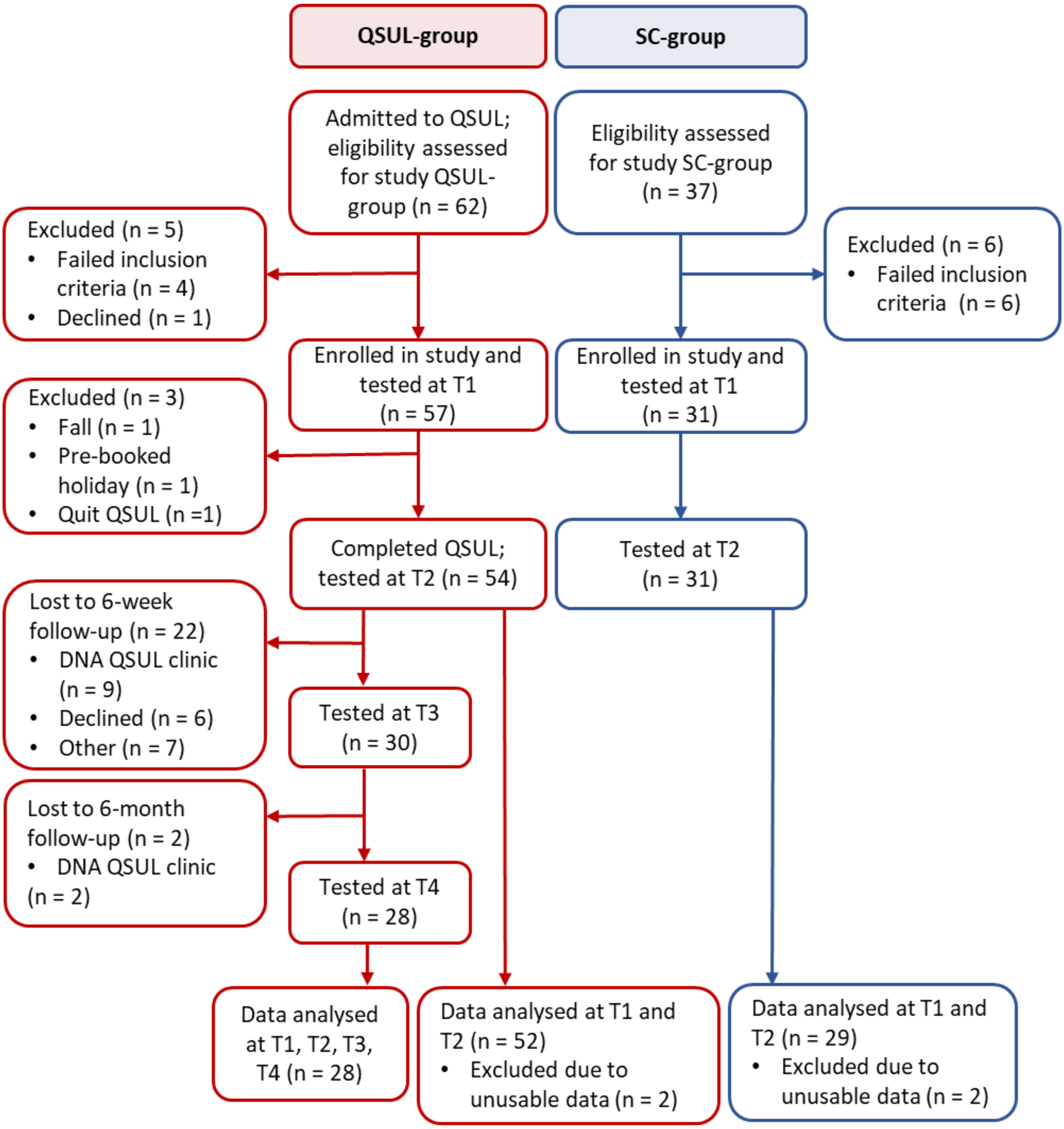


**Figure S1 CONSORT diagram illustrating patient recruitment to the study.**
All patients admitted to QSUL during the study period were assessed for study eligibility and included if they met the criteria and were willing to take part. The SC-group was recruited from the waiting list for QSUL, through local advertisement at the National Hospital for Neurology and Neurosurgery, Queen Square, online and via social media. A total of 81 patients, 52 QSUL-patients and 29 SC-patients, were included in the final two-timepoint analysis: change between T1 (0 weeks; admission to QSUL) and T2 (3 weeks; discharge from QSUL). In addition, follow-up analyses were performed on data from a subset of 28 QSUL-patients in a four-timepoint analysis: change between T1 and T2; change between T1 and T3 (6-week follow-up from QSUL); change between T1 and T4 (6-month follow-up from QSUL).

**Supplementary material: Control analyses**

Supplementary analyses were performed to investigate factors that might contribute to changes in kinematic measures.

**1. Do changes in impaired arm motor control reflect a change in speed-accuracy trade off?**
*Methods:* (A) It is important to check that kinematic improvement (especially accuracy) cannot simply be explained by a slower movement speed, where the speed-accuracy trade off (SAT) function would not changed. True improvement in motor control (assayed with kinematics) will be reflected in reduced SAT. We therefore calculated the SAT as the product of movement time and radial distance from target across all trials for each participant.

(B) In addition, the mean changes in movement time per trial were compared to (i) the mean change in radial distance from target and (ii) the mean change in the number of successful trials scored for each patient.

(C) Lastly, the mean ± standard deviation maximum velocity per trial were used to define the upper and lower maximum velocity parameters which encompass two-thirds of trials and excludes extremes at both high and low ends of the velocity profile. The kinematic analyses were repeated for trials fulfilling the velocity criteria $0.06 \leq$ max. velocity $\leq 0.11$ m/s only.

*Results:* (A) We compared Speed Accuracy Trade-Off (SAT) within and between QSUL and SC groups for T1 and T2. For the within group analysis, paired' Wilcoxon signed rank tests were used to compare SAT from T1 to T2. QSUL patients showed a significant improvement in SAT scores from T1 to T2 ($z = 2.60$, $p = 0.009$, $\eta 2 = 0.49$; Figure S2a), whereas no significant improvement was found for SC patients ($z = 0.67$ $p = 0.504$, $\eta 2 = 0.03$, Figure S2b). For the between group comparison, we compared the T1 to T2 SAT change for each group using an unpaired Wilcoxon rank sum test. The SAT improvement was greater for QSUL compared to SC groups ($z = 2.01$, $p = 0.04$, $\eta 2 = 0.58$, Figure S2c).

(B) Plots of change in movement time versus accuracy (radial distance from target) and change in successful trials (Supplementary Figures S2d and S2e) illustrate that the majority of patients improved in both speed and accuracy (dark green dots)).

(C) Furthermore, when the kinematic analyses were repeated for trials fulfilling the velocity criteria $0.06 \leq$ max. velocity $\leq 0.11$ m/s only, we still observed significant improvements in movement time, accuracy and smoothness (jerk) (Supplementary Figure S3).

However, all these results should be interpreted in the context that we did not systematically vary the movement speed during data collection and all participants were specifically instructed to complete each trial as quickly and accurately as possible.

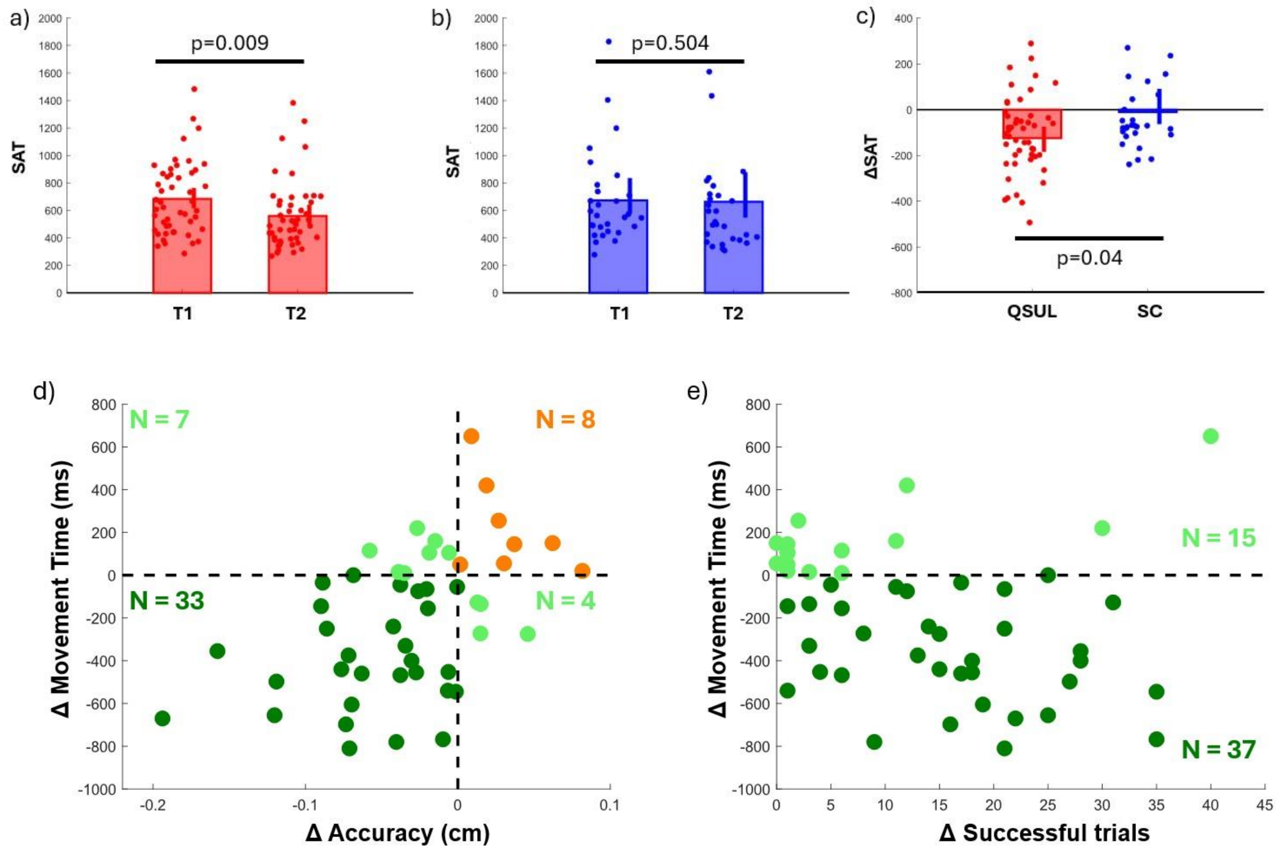


**Figure S2 Speed Accuracy Trade Off.** (a-c) Speed accuracy trade-off (SAT) performance for T1 and T2 for a) QSUL group; b) SC group; c) changes in SAT from T1-T2 for QSUL and SC groups. (d-e) Scatter plots depict changes from T1 to T2 in movement time versus (d) change in accuracy (radial distance from target) and (e) change in successful trials, all for the paretic arm of the QSUL group. Each dot represents one participant. Dark green dots indicate participants who improved in both metrics, while light green dots show those who improved in one metric. Orange dots represent patients who did not improve in either metric between T1 and T2. The mean changes are as follows: movement time (-206 ms), radial distance from target (-0.04 cm), successful trials (+12.3), all of which are in the direction of improvement.

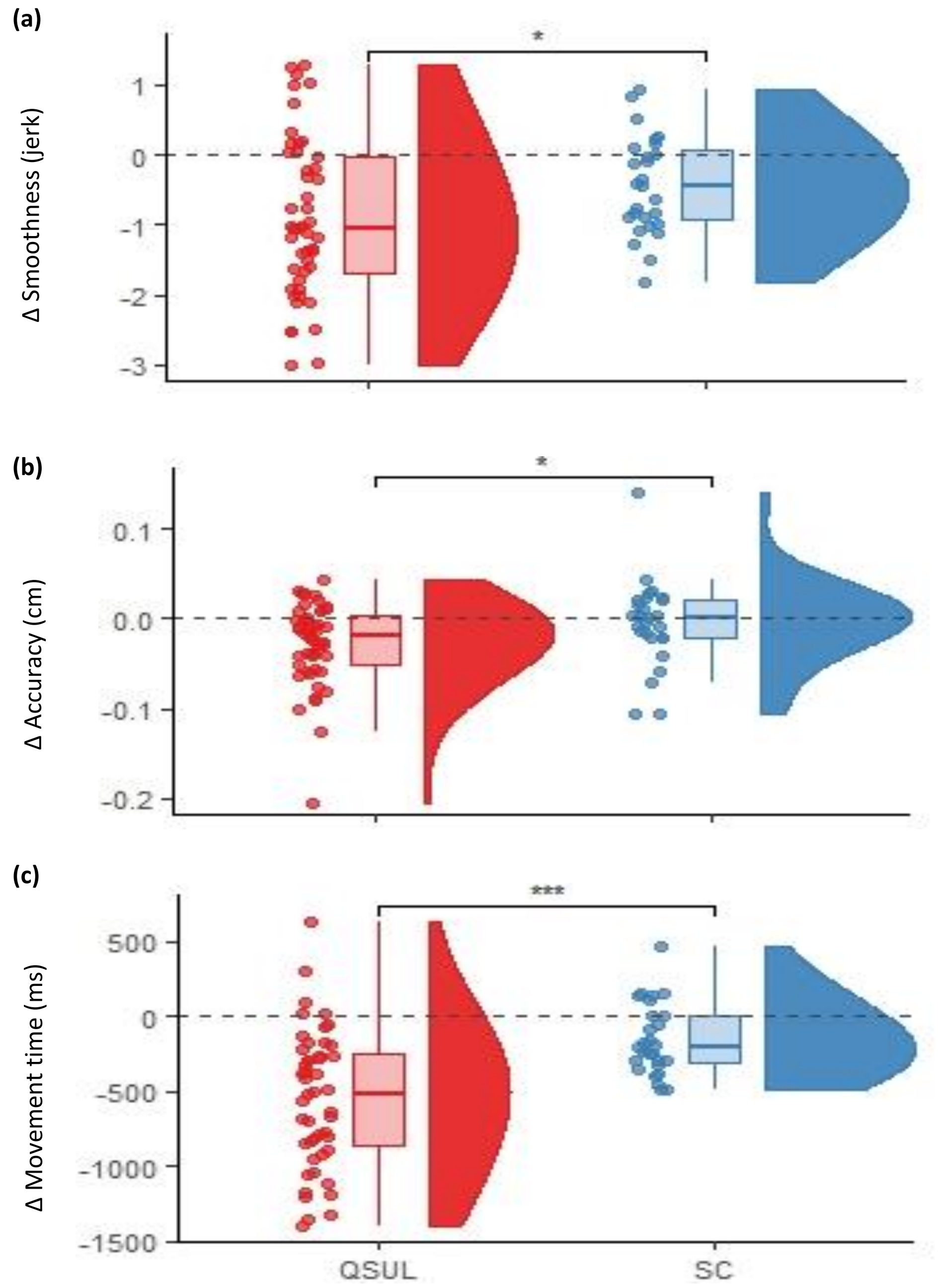


**Figure S3 Control analyses using velocity-stratification.**
The amount of change (Δ) from T1 to T2 in key kinematic parameters of whole movement trajectories for velocity-matched trials with 0.06 ≤ maximum velocity ≤ 0.11 m/s, performed by the impaired arm of the QSUL-group (shown in red), compared with the SC-group (shown in blue). The QSUL-group changed significantly more on **(a)** movement smoothness (jerk) **(b)** movement accuracy (radial distance from target, cm) and **(c)** movement time (ms), than the SC-group. The dashed grey line represents no change. The boxplots denote the median and interquartile range values. Differences in change scores were compared using 2-sample t-test with P values adjusted for multiple comparisons using the Bonferroni method *(P* ≤ 0.001 '***', *P* ≤ 0.01 '**', *P* ≤ 0.05 '*').

### 2. Are changes in impaired arm motor control independent of changes in active elbow ROM and arm strength?

*Methods:* While strength and motor control are dissociable in post-stroke motor recovery[1], we wanted to check that our observed improvements in movement quality were not simply a result of improved active ROM or strengthening at the elbow. Multiple linear regression mixed model analysis was performed to establish the extent to which any changes in kinematic measures could be accounted for by changes in active elbow ROM and/or elbow strength. Models were constructed with change in each of the four key kinematic parameters between T1 and T2 as the dependent variable in turn (i.e. movement smoothness (jerk), movement accuracy and movement time) and included changes in active ROM for both elbow flexion and extension, as well as biceps and triceps strength, as fixed effects with individual subject as a random effect. Explanatory variables were scaled to centre on a mean of zero with a standard deviation of one. Models were fitted using restricted maximum likelihood (REML) as the default parameter estimation criterion for linear mixed models. To determine the significance of the contribution made by each fixed effect, an alternative model without this fixed effect was constructed. Classical model comparison was then performed using an ANOVA test with the model objects as arguments to compare the fits of the alternative and null models, accepting a $P$ value of $< 0.05$ as significant. All multiple linear regression mixed model analyses were performed in R (version 3.6.1 (2019-07-05)) using the lme4 package.

*Results:* The models demonstrated that changes in active elbow ROM and elbow strength did not account for change in any kinematic measures ($P$ values for classical model comparison method all $> 0.05$). (Supplementary Table S1 Models A to C).

1. Xu J, Ejaz N, Hertler B, et al. Separable systems for recovery of finger strength and control after stroke. *J Neurophysiol*. 2017;118(2):1151-1163. doi:10.1152/jn.00123.2017

| **Model A (Δ_T1-T2)** | **Random Effects** | | | | **Fixed Effects** | | | | | | | | |
|---|---|---|---|---|---|---|---|---|---|---|---|---|---|
| Dependent variable | | Variance | % total variance | SD | | Estimate of variance (β) | % total variance | SE | t | $\chi^2$ | Df | p | Sig |
| Δ_Movement smoothness (Δ_jerk) | Subject (intercept) | 0.03 | 3.18 | 0.18 | Intercept | -0.01 | - | 0.04 | -0.15 | - | - | - | - |
| | Residual | 0.96 | - | 0.98 | Δ_Biceps strength | -0.99 x $10^{-2}$ | 0.79 | 0.04 | -0.24 | 0.06 | 1 | 0.81 | ns |
| | (No. obs 1853) | | | | Δ_Triceps Strength | -0.25 x $10^{-2}$ | 0.20 | 0.04 | -0.10 | 0.01 | 1 | 0.95 | ns |
| | | | | | Δ_Elbow flexion | -0.02 | 1.60 | 0.04 | -0.54 | 0.30 | 1 | 0.59 | ns |
| | | | | | Δ_Elbow extension | 0.02 | 1.60 | 0.04 | 0.66 | 0.43 | 1 | 0.51 | ns |

| **Model B (Δ_T1-T2)** | **Random Effects** | | | | **Fixed Effects** | | | | | | | | |
|---|---|---|---|---|---|---|---|---|---|---|---|---|---|
| Dependent variable | | Variance | % total variance | SD | | Estimate of variance (β) | % total variance | SE | t | $\chi^2$ | Df | p | Sig |
| Δ_Movement accuracy (Δ_radial dist. target, cm) | Subject (intercept) | 0.02 | 1.38 | 0.12 | Intercept | 0.76 x $10^{-3}$ | - | 0.03 | -0.03 | - | - | - | - |
| | Residual | 0.97 | - | 0.98 | Δ_Biceps strength | 0.03 | 2.34 | 0.03 | 0.76 | 0.58 | 1 | 0.45 | ns |
| | (No. obs 1853) | | | | Δ_Triceps Strength | -0.05 | 4.37 | 0.03 | -1.52 | 2.23 | 1 | 0.14 | ns |
| | | | | | Δ_Elbow flexion | -0.02 | 1.89 | 0.03 | -0.69 | 0.47 | 1 | 0.49 | ns |
| | | | | | Δ_Elbow extension | 0.04 | 3.31 | 0.03 | 1.23 | 1.47 | 1 | 0.23 | ns |

| **Model C (Δ_T1-T2)** | **Random Effects** | | | | **Fixed Effects** | | | | | | | | |
|---|---|---|---|---|---|---|---|---|---|---|---|---|---|
| Dependent variable | | Variance | % total variance | SD | | Estimate of variance (β) | % total variance | SE | t | $\chi^2$ | Df | p | Sig |
| Δ_Movement time (ms) | Subject (intercept) | 0.02 | 1.60 | 0.14 | Intercept | 0.06 x $10^{-2}$ | - | 0.03 | 0.22 | - | - | - | - |
| | Residual | 0.95 | - | 0.97 | Δ_Biceps strength | 0.01 | 0.90 | 0.03 | 0.31 | 0.10 | 1 | 0.76 | ns |
| | (No. obs 1853) | | | | Δ_Triceps Strength | -0.07 | 5.80 | 0.03 | -2.13 | 4.35 | 1 | 0.07 | ns |
| | | | | | Δ_Elbow flexion | 0.11 | 8.70 | 0.03 | 3.37 | 7.33 | 1 | 0.06 | ns |
| | | | | | Δ_Elbow extension | 0.08 | 6.40 | 0.03 | 2.50 | 5.6 | 1 | 0.12 | ns |

**Table S1** Multiple linear regression mixed models for change in measures between T1-T2 in the impaired arm of the QSUL group (n = 52 patients). Dependent variables: Model A – Movement smoothness (jerk); Model B – Movement accuracy; Model C – Movement time.

**3. Are changes in impaired arm motor control due to improved movement execution or feedback control?**

*Methods:* To test whether improvements in motor control came about due to enhanced execution of the planned movement rather than improved ability to make online corrections through enhanced feedback control[2,3] our analyses were repeated using only the initial outward submovement of the whole movement trajectory (so that online corrections could not account for improvements), defined as the movement until the first minimum velocity (<0.005 m/s).

*Results:* From T1 to T2, there were improvements in the QSUL-group compared to SC-group for (a) the proportion of the whole movement trajectory time taken up with the primary outward submovement; (b) movement smoothness (jerk) and (c) movement accuracy (Supplementary Figure S4). These results suggest improved movement execution rather than just getting better at online compensatory corrections.

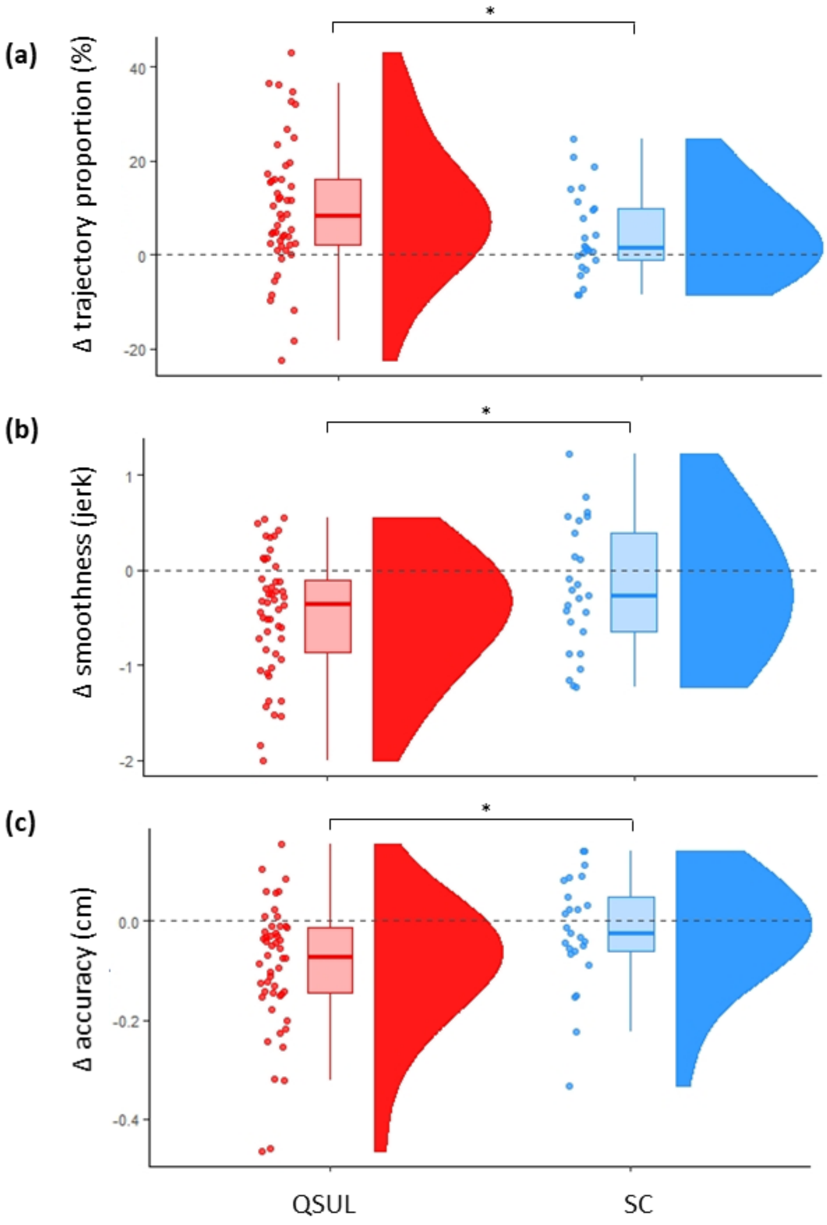

**Figure S4 Initial outward submovement trajectory analysis.**
The amount of change (Δ) from T1 to T2 in key kinematic parameters of the primary outward submovement trajectories for velocity-matched trials with $0.06 \leq$ maximum velocity $\leq 0.11$ m/s, performed by the impaired arm of the QSUL-group, compared with the SC-group. The QSUL-group changed significantly more on key kinematic measures than the SC-group, suggesting improved movement execution as opposed to enhanced feedback control. **(a)** change (Δ) in primary outward submovement as a proportion of the whole movement trajectory (the higher the proportion, the better the movement quality), **(b)** change in submovement smoothness (jerk) and **(c)** change in submovement accuracy (cm). The primary outward submovement of the movement trajectory was defined as the movement up until the first minimum velocity (<0.005 m/s). The dashed grey line represents no change. The boxplots denote the median and interquartile range values. Differences in change scores were compared using 2-sample 2-tailed t-test with P values adjusted for multiple comparisons using the Bonferroni method (all $P < 0.05$).

2. Shmuelof L, Krakauer JW, Mazzoni P. How is a motor skill learned? Change and invariance at the levels of task success and trajectory control. J Neurophysiol. 2012;108(2):578-594. doi:10.1152/jn.00856.2011

3. Pratt J, Chasteen AL, Abrams RA. Rapid aimed limb movements: age differences and practice effects in component submovements. Psychol Aging. 1994;9(2):325-334. doi:10.1037//0882-7974.9.2.325

### 4. Are changes in impaired arm motor control related to online learning during kinematic testing?

Methods: To assess whether changes in performance of the paretic arm were due to online learning, we calculated average performance at the beginning (Early – first 50% of trials) and end (Late – second 50% of trials) of each assessment day (T1 and T2). We conducted paired Wilcoxon signed rank tests comparing (i) T1 Early to T1 Late, (ii) T1 Late to T2 Early; and (iii) T2 Late to T2 Early) for each of speed-accuracy trade off (SAT), movement time, accuracy (radial distance from target) and smoothness (jerk), for each group. We also performed a between-group comparison for each outcome measure to assess whether QSUL and SC patients showed differences in online learning between T1 Early and T1 Late, using an unpaired Wilcoxon rank sum test.

Results: There was no evidence of within session online learning for either the QSUL-group or SC-group for each of speed-accuracy trade off (SAT), movement time, accuracy (radial distance from target) and smoothness (jerk). However, there were significant improvements from T1 Late to T2 Early for all of these measures in the QSUL-group, but not SC-group (Supplementary Figures S5a and b). Direct comparison of within T1 changes, confirmed no differences between the QSUL-group and SC-group group (Supplementary Figure S5c).

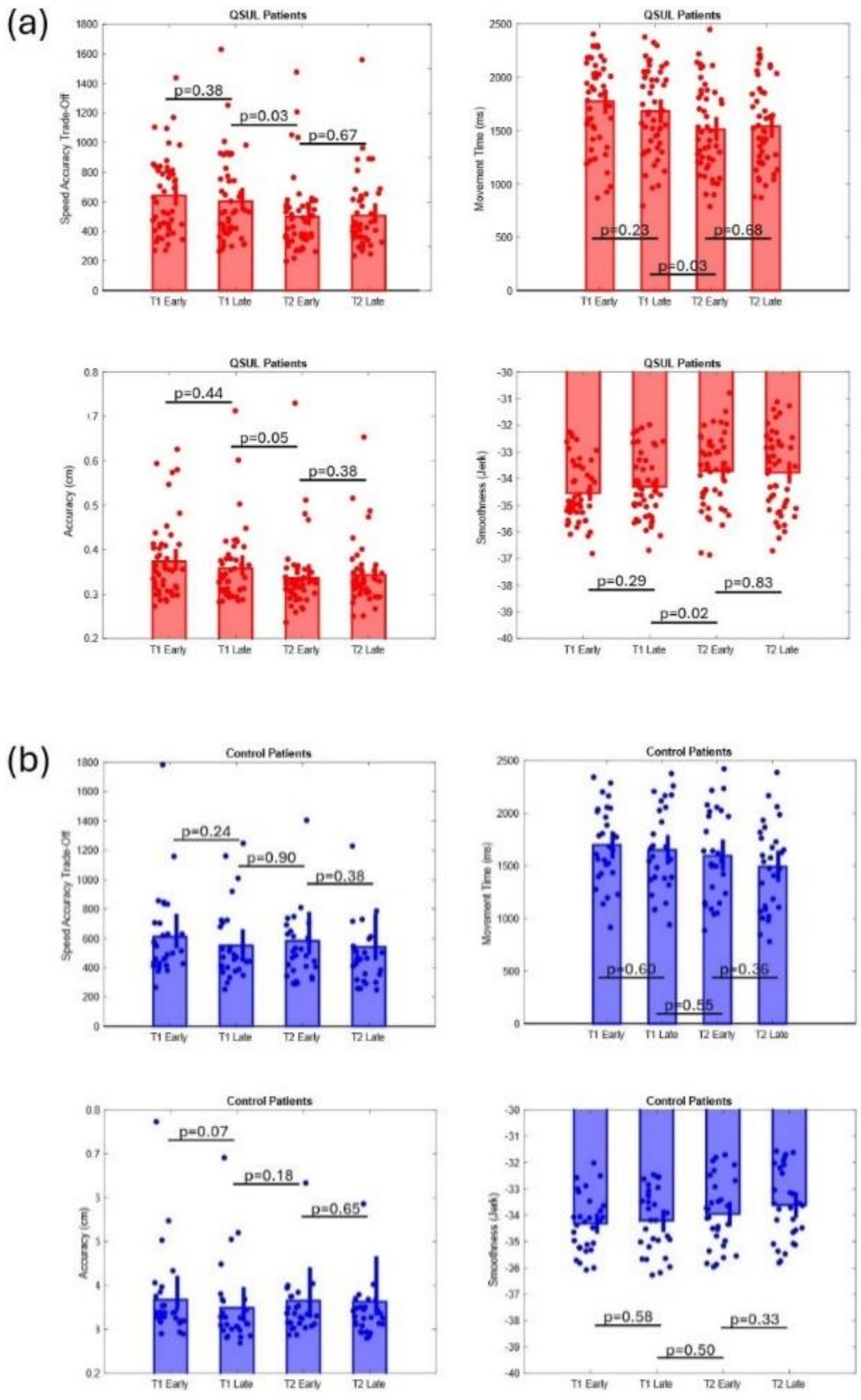
(a)
QSUL Patients
Speed Accuracy Trade-Off
p=0.38
p=0.03
p=0.67
Movement Time (ms)
p=0.23
p=0.68
p=0.03
Accuracy (cm)
p=0.44
p=0.05
p=0.38
Smoothness (Jerk)
p=0.29
p=0.83
p=0.02
T1 Early
T1 Late
T2 Early
T2 Late
(b)
Control Patients
Speed Accuracy Trade-Off
p=0.24
p=0.90
p=0.38
Movement Time (ms)
p=0.60
p=0.36
p=0.55
Accuracy (cm)
p=0.07
p=0.18
p=0.65
Smoothness (Jerk)
p=0.58
p=0.33
p=0.50

(c)

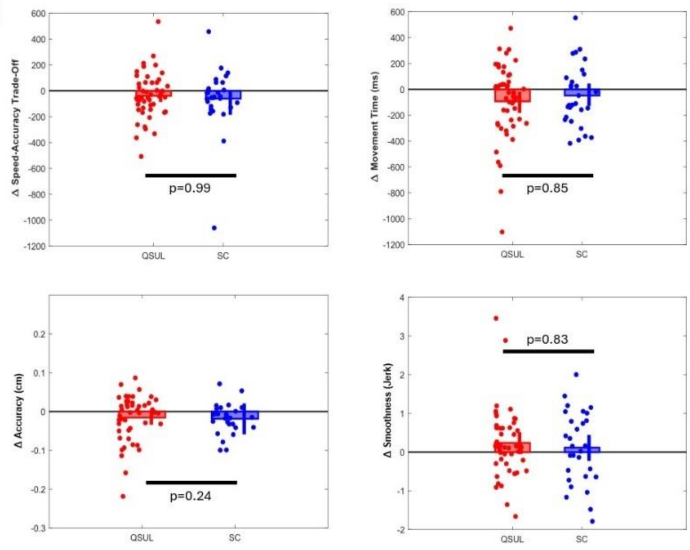


**Figure S5 Assessment of within and between session learning**

To assess whether changes in performance of the paretic arm were due to online learning, we calculated average performance at the beginning (Early – first 50% of trials) and end (Late – second 50% of trials) of each assessment day (T1 and T2). We conducted 'paired' Wilcoxon signed rank tests comparing (i) T1 Early to T1 Late, (ii) T1 Late to T2 Early; and 'unpaired 'Wilcoxon' rank sum tests comparing (iii) T2 Late to T2 Early) for each of speed-accuracy trade off (SAT), movement time, accuracy (radial distance from target) and smoothness (jerk) for each group.

(a) QSUL-group: There was no evidence of <u>within</u> session online learning for the QSUL group:

SAT (T1 Early vs. T1 Late: z = 0.88, p = 0.38, η2 = 0.14); T2 Early vs. T2 Late: z = -0.43, p = 0.67, η2 = 0.03); Movement Time (T1 Early vs. T1 Late: z = 1.19, p = 0.23, η2= 0.23; T2 Early vs. T2 Late: z = -0.42, p = 0.68, η2 = 0.07); Accuracy (radial distance from target) (T1 Early vs. T1 Late: z = 0.77, p = 0.44, η2 = 0.18; T2 Early vs. T2 Late: z = -0.89, p = 0.38, η2 = 0.09); or Smoothness (jerk) (T1 Early vs. T1 Late: z = -1.10, p = 0.29, η2 = 0.21); T2 Early vs. T2 Late: z = 0.21, p = 0.83, η2 = 0.05).

However, there were significant improvements from T1 Late to T2 Early in SAT (z = 2.13, p = 0.03, η2= 0.40); Movement Time (z = 2.14, p = 0.03, η2 = 0.42); Accuracy (radial distance from target) (z = 1.92, p = 0.05, η2 = 0.32); and Smoothness (jerk) (z = -2.29, p = 0.02, η2 = 0.47).

(b) SC-group: There was no evidence of within session online learning or improvements from T1 Late to T2 Early for the SC group:

SAT: (T1 Early vs. T1 Late: z = 1.18, p = 0.24, η2 = 0.22; T2 Early vs. T2 Late: z = 0.90, p = 0.38, η2 = 0.11; T1 Late vs T2 Early: z = -0.12, p = 0.90, η2 = 0.10); Movement Time: (T1 Early vs. T1 Late: z = 0.52, p = 0.60, η2 = 0.13; T2 Early vs. T2 Late: z = 0.91, p = 0.36, η2 = 0.25; T1 Late vs T2 Early: z = 0.60, p = 0.55, η2 = 0.13); Smoothness (jerk): (T1 Early vs. T1 Late: z = -0.56, p = 0.58, η2 = 0.10; T2 Early vs. T2 Late: z = -0.98, p = 0.33, η2 = 0.27; T1 Late vs T2 Early: z = 0.67, p = 0.50 η2 = 0.20); Accuracy (radial distance from target): (T1 Early vs. T1 Late: z = 1.79, p = 0.07, η2 = 0.20; T2 Early vs. T2 Late: z = 0.45, p = 0.65, η2 = 0.02; T1 Late vs T2 Early: z = -1.33, p = 0.18, η2 = 0.16).

(c) Between group comparison: We further ran a between-group comparison for each outcome measure to assess whether QSUL and SC patients showed differences in online learning between T1 Early and T1 Late, using an 'unpaired' Wilcoxon rank sum test. Results confirmed that there were no significant differences between groups (SAT: z = -0.01, p = 0.99, η2 = 0.10; Movement Time: z = 0.19, p = 0.85, η2 = 0.16; Accuracy: z = -1.18, p = 0.24, η2 = 0.04; Smoothness (jerk): z = -0.21, p = 0.83 η2 = 0.14).

**Supplementary material: Control comparisons involving less impaired ('normal') arms**

*Methods:* To ensure that QSUL-group impaired arm changes were attributable to the therapeutic intervention targeting that arm, rather than a non-specific effect of performing the task several times, we performed two further sets of analyses comparing the QSUL-group less impaired ('normal') arm to (i) the QSUL-group impaired arm, and (ii) the SC-group less impaired ('normal') arm. Note that during the rehabilitation intervention, treatment incorporates both arms to a certain extent, and so we might expect to see some small improvements in the less impaired ('normal') arm.

*Results:* Firstly, changes in elbow strength, elbow active ROM and kinematics were all greater in QSUL-group impaired arm compared to less impaired ('normal') arm (Table S2). Secondly, only biceps and triceps strength improved more in less impaired ('normal') arm of the QSUL-group compared to the SC-group, with no differences in active elbow ROM or kinematic measures (Table S3). All comparisons shown in Figure S6.

Taken together, these results suggest that the changes we report for the QSUL-group impaired arm that was the main focus of treatment were unlikely to be driven by a non-specific learning effect, such as repeating the kinematic task several times.

**Table S2** Change in clinical and kinematic measures between T1 and T2 for the impaired compared to the less impaired ('normal') arms of the QSUL-group (n = 52).

| Change (Δ) T1-T2 | QSUL-group IMPAIRED ARM | QSUL-group LESS IMPAIRED ('NORMAL') ARM | ANOVA[a] | |
|---|---|---|---|---|
| | Med (1/4-3/4) | Med (1/4-3/4) | *F*(1,79) | p |
| **Clinical Measures** | | | | |
| Δ_Biceps strength (kgf) | 0.80 (0.24-2.08) | 0.28 (0.00-0.91) | 3.12 | 0.04 |
| Δ_Triceps strength (kgf) | 0.60 (0.18-2.35) | 0.22 (-0.01-0.83) | 3.36 | 0.04 |
| Δ_Elbow flexion (deg) | 2.00 (0.00-5.00) | 0.00 (0.00-0.00) | 13.77 | $0.34 \times 10^{-3}$ |
| Δ_Elbow extension (deg) | 0.00 (-9.25-0.00) | 0.00 (0.00-0.00) | 20.41 | $0.18 \times 10^{-4}$ |
| **Kinematic measures** | | | | |
| Δ_Movement smoothness (jerk) | -0.68 (-1.36-(-0.11)) | -0.22 (-0.66-0.03) | 10.10 | $0.20 \times 10^{-2}$ |
| Δ_Movement accuracy (radial dist. cm) | -0.02 (-0.05-0.00) | 0.00 (-0.01-0.01) | 11.56 | $0.97 \times 10^{-3}$ |
| Δ_Movement time (ms) | -227.50 (-455.63-(-10.00)) | -46.25 (-145.00-11.88) | 8.98 | $0.35 \times 10^{-2}$ |

[a]Changes in clinical and kinematic measures between T1 and T2for each arm of the QSUL-group were compared using 2x2 mixed ANOVAs for significant group x timepoint interactions. Med: median value, 1/4: 25th percentile, 3/4: 75th percentile.

**Table S3** Change in clinical and kinematic measures between T1 and T2 for the less impaired ('normal') arms of the QSUL-group (n = 52) compared to the SC-group (n = 29).

| Change (Δ) T1-T2 | QSUL-group | SC-group | ANOVA[a] | |
|---|---|---|---|---|
| **LESS IMPAIRED ('NORMAL') ARM** | **Med (1/4-3/4)** | **Med (1/4-3/4)** | ***F*(1,73)** | **p** |
| **Clinical Measures** | | | | |
| Δ_Biceps strength (kgf) | 0.28 (0.00-0.91) | 0.00 (-0.18-0.28) | 7.82 | 0.01 |
| Δ_Triceps strength (kgf) | 0.22 (-0.01-0.83) | -0.10 (-0.27-0.050 | 5.94 | 0.02 |
| Δ_Elbow flexion (deg) | 0.00 (0.00-0.00) | 0.00 (0.00-0.00) | 2.68 | 0.12 |
| Δ_Elbow extension (deg) | 0.00 (0.00-0.00) | 0.00 (0.00-0.00) | 1.14 | 0.29 |
| **Kinematic measures** | | | | |
| Δ_Movement smoothness (jerk) | -0.22 (-0.66-0.03) | -0.17 (-0.60-0.11) | 0.28 | 0.60 |
| Δ_Movement accuracy (radial dist. cm) | 0.00 (-0.01-0.01) | 0.00 (-0.01-0.02) | 0.85 | 0.36 |
| Δ_Movement time (ms) | -46.25 (-145.00-11.88) | -22.50 (-170.00-86.25) | 0.26 | 0.61 |

[a]Changes in clinical and kinematic measures between T1 and T2 in the QSUL-group and SC-group were compared using 2x2 mixed ANOVAs for significant group x timepoint interactions. Med: median value, 1/4: 25$^{th}$ percentile, 3/4: 75$^{th}$ percentile.

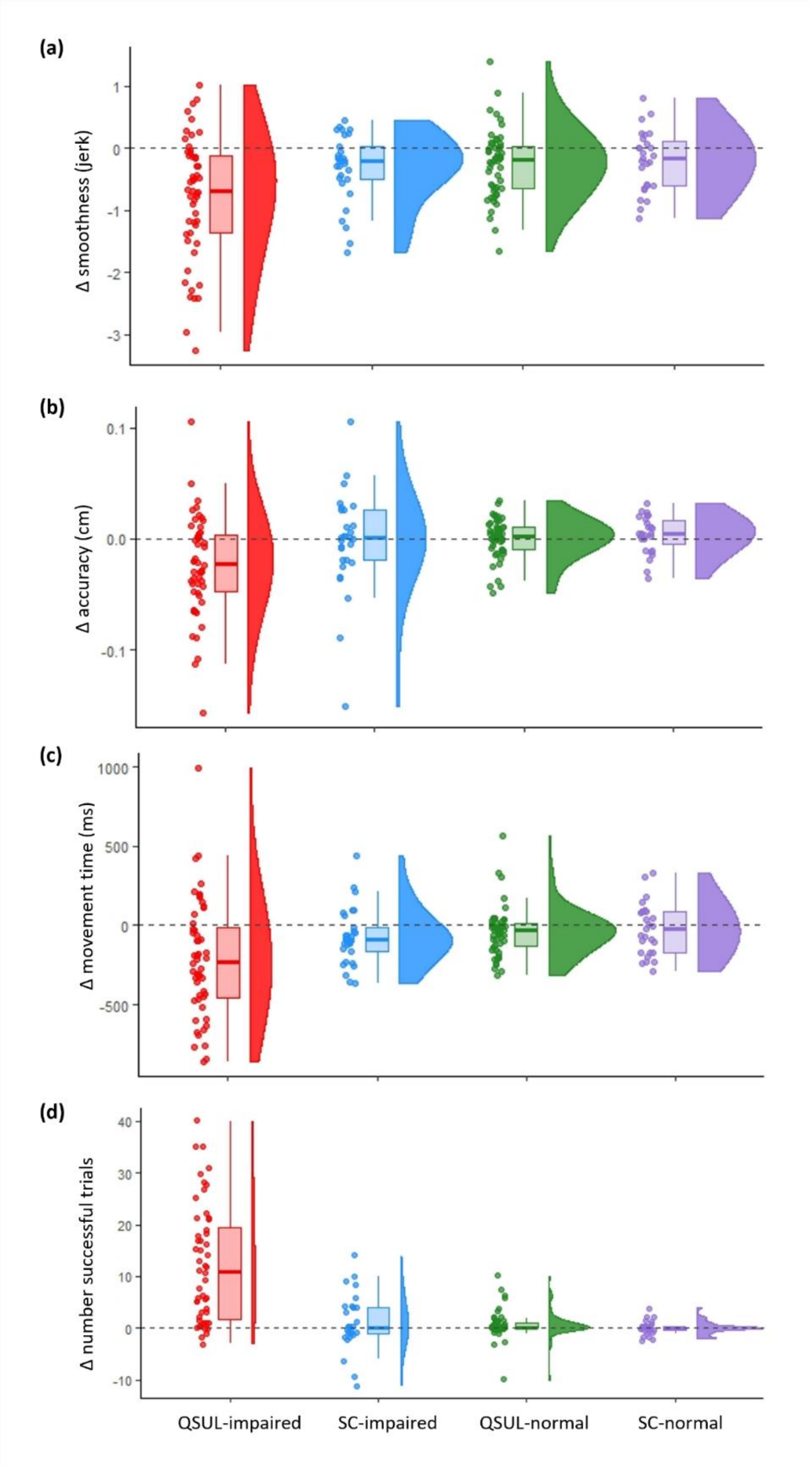


**Figure S6 Change (Δ) in key kinematic measures for the impaired and less impaired arms of the QSUL-group (n = 52), compared to the impaired and less impaired arms of the SC-group (n = 29) from T1 to T2. a)** Movement smoothness (jerk), **b)** movement accuracy and **c)** movement time. Improvement in each of these parameters is indicated by a reduction in the measure between timepoints. d) Number of successful trials: improvement is indicated by an increase in this measure. The dashed grey line represents no change. The boxplots denote the median and interquartile range values. The 1/2 violin plots illustrate the distribution of the data. QSUL-group impaired arm is shown in red, SC-group impaired arm is shown in blue, QSUL-group less impaired arm is shown in green, SC-group less impaired arm is shown in lilac
